# Beyond Accuracy: An Efficiency- and Safety-Aware Framework for Evaluating Clinical AI with Large Language Models

**DOI:** 10.1101/2025.10.14.25338039

**Authors:** Nazar Zaki, Amal Akor, Salahdein Aburuz, Sham ZainAlAbdin

## Abstract

**Background:** Large language models (LLMs) demonstrate strong performance on medical reasoning tasks, but current evaluation approaches focus primarily on accuracy, neglecting the efficiency–safety trade-offs critical for real-world clinical utility.

**Methods:** We developed and validated the Clinical Value Density (CVD) framework, a novel metric quantifying clinical utility per unit of cognitive resource consumed. Six state-of-the-art LLMs (GPT-4o, Gemini-2.5, Claude-Sonnet-4, Grok-3, DeepSeek-R1, and Kimi-K2) were evaluated across 60 authentic clinical pharmacology scenarios derived from UAE healthcare practice and benchmarked against board-certified clinical pharmacist responses. Performance was assessed across efficiency, semantic similarity, safety, relevance, consistency, and conciseness, with triangulated validation against clinician preference and task efficiency.

**Results:** Traditional metrics (BLEU: 0.003–0.024; ROUGE-L: 0.079–0.166) failed to reflect clinical utility, while the CVD framework exposed critical efficiency–safety trade-offs. GPT-4o achieved the highest normalized CVD (0.475), delivering fourfold efficiency gains over pharmacists (41 vs 178 tokens) but with moderate safety (0.437), requiring supervised deployment. Grok-3 and Gemini-2.5 led in safety (0.605 and 0.542) but at the cost of efficiency. DeepSeek-R1 and Kimi-K2 produced unsafe brevity–accuracy trade-offs, generating concise but clinically inaccurate responses. Comparative validation revealed strong alignment between CVD and task efficiency (r = 0.924), but divergence from clinician preference (r = –0.845), reflecting a bias toward verbose outputs.

**Conclusions:** Current LLMs cannot reliably perform clinical functions autonomously. Instead, they are best positioned for AI-assisted, supervised integration, where efficiency and safety are balanced under professional oversight. The CVD framework provides a potentially useful for regulatory evaluation for quantifying deployment readiness, aligning AI evaluation with the real-world constraints of cognitive load, time, and patient safety. Future research should extend CVD across specialties, scale validation datasets, and conduct real-time workflow trials to establish specialty-specific safety thresholds for eventual autonomy.

**Author Summary:** Large language models (LLMs) are impressively accurate on medical reasoning benchmarks, yet clinical work is not a quiz, it is a race against time under strict safety constraints. We introduce Clinical Value Density (CVD), a simple yet powerful metric that captures clinical utility per unit of cognitive cost (i.e., how much useful care an answer delivers for the effort it imposes on clinicians). In a head-to-head evaluation of six state-of-the-art LLMs across 60 authentic clinical pharmacology scenarios benchmarked against board-certified pharmacists, traditional metrics (BLEU/ROUGE) barely moved, while CVD exposed the true efficiency–safety trade-offs that determine bedside usefulness. For example, GPT-4o delivered the highest normalized CVD (0.475) with ∼4× fewer tokens than pharmacists (≈41 vs 178), but only moderate safety (0.437) appropriate for supervised use, not autonomy. Conversely, Grok-3 and Gemini-2.5 were safest but less efficient; DeepSeek-R1 and Kimi-K2 risked unsafe brevity– accuracy trade-offs. Crucially, CVD strongly tracked task efficiency (r=0.924) yet diverged from clinician preference (r=–0.845), revealing a bias toward overly verbose answers that feel reassuring but slow care.

## 1. Introduction

Artificial intelligence (AI) has emerged as one of the most transformative forces in healthcare, with large language models (LLMs) demonstrating strong performance in diagnostic support, treatment planning, and clinical decision-making (Singhal et al., 2023; Thirunavukarasu et al., 2023; Lee et al., 2023). Recent foundation models have achieved human-level performance on medical examinations and shown competency in complex reasoning tasks (Nori et al., 2023; Liévin et al., 2024; Yang et al., 2022). Yet the central question remains unresolved: can these systems safely perform clinical functions autonomously, or do they require supervised deployment under human oversight?

The translation of AI success from laboratory benchmarks to clinical practice is limited by cognitive and temporal constraints. For example, primary care physicians average only 13–16 minutes per patient encounter (Irving et al., 2017). Emergency clinicians often manage multiple cases at once (Horwitz et al., 2019). In pharmacy practice, drug information consultations frequently exceed 11 minutes per query (Malone et al., 2018). Under such conditions, verbose or poorly structured outputs can be as problematic as inaccurate ones. Systematic reviews confirm that information overload frequently leads to abandonment of decision support tools, regardless of accuracy (Johnson et al., 2005). Challenges are especially acute in rapidly expanding healthcare systems such as the United Arab Emirates (UAE), where high burdens of chronic and complex diseases strain limited resources (UAE Ministry of Health and Prevention, 2024). Within this environment, clinical pharmacology provides a critical testbed, as board-certified pharmacists must deliver accurate, evidence-based responses under severe time pressures (American College of Clinical Pharmacy, 2008).

Traditional evaluation methods such as BLEU, ROUGE, and embedding-based metrics capture factual accuracy but neglect the efficiency–quality trade-offs central to clinical adoption (Papineni et al., 2002; Lin, 2004; Reimers & Gurevych, 2019). Studies show these metrics consistently favor verbose outputs, even though longer responses increase cognitive load and reduce adoption (Chen et al., 2023; Berner et al., 1994; Sittig et al., 2008; Patel et al., 2008). Martinez et al. (2008) further demonstrated that general-purpose Natural language Processing (NLP) measures often fail to predict clinical utility, as they systematically ignore efficiency constraints. Since healthcare fundamentally operates as a problem of cognitive resource allocation, with practitioners needing to filter relevant information quickly (Norman & Bobrow, 1975; Wickens, 2002), AI systems must optimize information density to support rather than burden workflows (Miller, 1956; Chase & Simon, 1973; Ericsson & Kintsch, 1995). Tools requiring excessive cognitive processing are often abandoned, even when accurate (Davis et al., 1989). Clinical pharmacology exemplifies this demand, as pharmacists must appraise literature, synthesize evidence, and communicate responses within narrow timeframes (American Society of Health-System Pharmacists, 2015).

The rapid advancement of LLMs amplifies the need for efficiency-integrated evaluation frameworks. Benchmarking studies reveal substantial variation in model response styles, with some producing concise summaries and others verbose outputs (OpenAI, 2023; Anil et al., 2023; Anthropic, 2023). Current evaluation approaches provide little guidance on how such characteristics align with real-world clinical settings, where efficiency requirements differ by specialty. In pharmacology, delays or inaccuracies in drug information can directly compromise patient safety (Bond & Raehl, 2007; Kaboli et al., 2006). Studies show that efficiency strongly correlates with consultation throughput and access to pharmaceutical expertise (Rough et al., 1982). However, domain-specific evaluation approaches remain rare (Lee et al., 2020). Fewer than a dozen studies have addressed this issue, and most of them focused narrowly on response time (Roberts et al., 2017; Wang et al., 2018).

Recent advances in NLP, including semantic embedding and domain-specific similarity metrics (Devlin et al., 2019; Kenton & Toutanova, 2019), offer a foundation for efficiency-aware evaluation, but these methods have not yet been adapted to the realities of clinical workflows, safety requirements, or professional standards.

Clinical pharmacology provides an ideal validation domain, given its well-established professional standards, structured evidence synthesis processes, and measurable consultation outcomes (Koda-Kimble et al., 2018; Nathan et al., 2007). Drug information services typically demand comprehensive yet timely responses, averaging more than 11 minutes for complex queries (Rosenberg et al., 2004). This creates authentic conditions to evaluate whether AI systems can balance depth with speed. Regional contexts such as the UAE healthcare system add further validation by embedding evaluation frameworks in authentic epidemiological and therapeutic environments (Alsairafi et al., 2020), ensuring both methodological rigor and cultural relevance.

Against this background, the present study addresses the critical question of clinical AI autonomy: Can current LLMs perform clinical functions autonomously, or do they require ongoing human supervision? We propose and validate a Clinical Value Density (CVD) framework, designed to quantify semantic quality per unit of cognitive load and thereby capture the efficiency quality trade-offs central to clinical practice. The study pursues the following objectives.

### 1.1. Study Objectives

**Primary objective:**

To develop and validate the Clinical Value Density (CVD) framework to enable systematic comparison of LLMs and determine deployment readiness and supervision requirements.

**Secondary Objectives:**

1. To compare six state-of-the-art LLMs (GPT-4o (OpenAI, 2024), Gemini-2.5 (Kavukcuoglu, 2025), Claude-Sonnet-4 (Anthropic, 2025), Grok-3 (xAI, 2025), DeepSeek-R1 (DeepSeek-AI *et al*., 2025), and Kimi-K2 (Kimi Team, 2025)) using the CVD framework across 60 UAE-derived clinical pharmacology scenarios validated by board-certified pharmacists.
2. To analyze efficiency–quality trade-offs and error patterns, providing evidence-based guidance on deployment strategies (autonomous, supervised, or AI-assisted).
3. To integrate safety scoring into the framework in order to identify dangerous efficiency–accuracy trade-offs that could compromise patient outcomes.
4. To benchmark CVD against conventional NLP metrics and professional standards, demonstrating its capacity to distinguish between deployment-ready and supervision-requiring systems.

This research contributes to clinical AI evaluation in three key ways: first, by integrating efficiency and safety into a quantitative framework validated against authentic workflows rather than artificial benchmarks; second, by embedding safety considerations to identify trade-offs that could endanger patients and mandate supervised deployment; and third, by providing comparative benchmarking of contemporary LLMs across authentic scenarios, offering actionable evidence for determining deployment thresholds. By focusing on clinical pharmacology where delayed or inaccurate drug information can have immediate consequences, this study delivers practical insights into the supervision needs of AI in real-world practice. From a methodological perspective, it advances evaluation science by moving beyond accuracy-focused metrics toward comprehensive frameworks that explicitly assess deployment readiness, supervision requirements, and safety risks, with regional validation ensuring global applicability.

## 2. Experimental Work and Results

### 2.1. Experimental Setup

A total of 60 clinical pharmacology scenarios were evaluated across six LLMs, reflecting the most prevalent diseases in the United Arab Emirates. Case distribution was balanced across cognitive demand levels: 23 low-complexity (38.3%), 17 intermediate-complexity (28.3%), and 20 high-complexity cases (33.3%). Disease categories included hypertension, diabetes, breast cancer, autism spectrum disorders, and infectious diseases, aligning with regional epidemiological patterns while remaining relevant to global healthcare contexts. Expert validation confirmed that low-complexity scenarios required retrieval from tertiary sources, intermediate cases demanded secondary literature synthesis, and high-complexity queries necessitated primary research appraisal and patient-centered reasoning. This stratification provided authentic representation of the decision-making spectrum encountered in clinical pharmacy practice.

The CVD framework revealed marked variation in performance across the six LLMs. Response length differed by nearly four-fold, from 41.0 tokens (GPT-4o) to 171.4 tokens (Claude-Sonnet-4), with direct implications for cognitive load. In contrast, clinical pharmacists required an average of 178.1 tokens and 11 minutes per consultation, providing realistic professional benchmarks.

Responses were preprocessed, tokenized, and analyzed using a multi-dimensional evaluation suite comprising:

- Traditional NLP metrics (BLEU, ROUGE-L, factuality).
- CVD metrics (raw and normalized semantic value per token).
- Clinical safety scoring (accuracy-weighted error categorization).
- Relevance density (domain keyword weighting).
- Conciseness quality (short, medium, long stratification with excellence bonuses).

Statistical validation employed paired t-tests with Holm-Bonferroni correction, effect size estimation (Cohen’s d), and cross-correlation analyses with traditional metrics.

### 2.2. Model Performance on Traditional and Baseline Metrics

To provide a comprehensive baseline, we compared six state-of-the-art large language models against board-certified clinical pharmacists across multiple evaluation dimensions. In addition to conventional NLP measures (BLEU, ROUGE-L, factuality), we report semantic similarity to clinician references, average response length (tokens), and response generation time **(Table 1).** Clinical pharmacist consultation time and response length are included for context, establishing authentic professional benchmarks.

**Table 1:** Comparative Performance of the LLMs and Clinical Pharmacists on Traditional Metrics and Response Characteristics.

| Model | Avg Tokens | Semantic Similarity | BLEU Score | ROUGE-L | Factuality Score | Response Time (sec) |
| --- | --- | --- | --- | --- | --- | --- |
| GPT-4o | 41.0 | 0.656 | 0.006 | 0.111 | 0.106 | 47.0 (±4.3) |
| Gemini-2.5 | 86.4 | 0.733 | 0.024 | 0.166 | 0.216 | 52.1 (±6.2) |
| Claude-Sonnet-4 | 171.4 | 0.703 | 0.013 | 0.121 | 0.197 | 68.3 (±8.1) |
| Grok-3 | 94.7 | **0.739** | 0.016 | 0.139 | 0.173 | 58.7 (±7.4) |
| DeepSeek-R1 | 44.6 | 0.502 | 0.003 | 0.079 | 0.051 | **43.2 (±5.1)** |
| Kimi-K2 | 50.9 | 0.533 | 0.003 | 0.099 | 0.067 | 45.8 (±4.9) |
| Clinical Pharmacists | 178.1 | --- | --- | --- | --- | 675.0 (±81.0) |

The expanded comparison highlights several critical patterns. First, semantic similarity to clinical references varied considerably, with Grok-3 (0.739) and Gemini-2.5 (0.733) outperforming GPT-4o (0.656), while DeepSeek-R1 (0.502) and Kimi-K2 (0.533) lagged behind. However, high semantic similarity did not translate into efficiency, as Claude-Sonnet-4 required 171.4 tokens per response over four times GPT-4o’s 41.0 tokens demonstrating verbosity as a barrier to clinical integration. Second, traditional metrics remained uniformly low (BLEU ≤ 0.024, ROUGE-L ≤ 0.166), reinforcing that surface-level lexical overlap does not adequately reflect clinical utility. For example, Gemini-2.5 achieved the highest factuality score (0.216) yet required twice the response length of GPT-4o, illustrating the inefficiency of verbose outputs despite high lexical recall. Third, response times demonstrated that LLMs operate orders of magnitude faster than clinicians. Even the slowest model (Claude-Sonnet-4, 68.3 seconds) was nearly ten times faster than the pharmacist benchmark (675 seconds). GPT-4o, with the lowest average token count and a 47-second response time, achieved the most favorable balance between speed and conciseness.

Moreover, the clinical pharmacist baseline emphasizes the cognitive and temporal intensity of professional consultations: 178 tokens on average, produced over 11 minutes. This establishes a realistic benchmark for cognitive load against which AI efficiency gains can be measured, while reminding that professional responses integrate current literature, nuanced reasoning, and safety considerations beyond what token counts capture. Collectively, these findings reveal that traditional metrics underestimate the clinical promise of LLMs, while raw efficiency indicators highlight models like GPT-4o as potential workflow enhancers when used under professional supervision.

While conventional metrics provided limited insight, the CVD framework exposed efficiency–quality trade-offs that remain invisible to traditional evaluation. **Figure 1** plots semantic similarity against CVD scores across all six models, with bubble size representing inverse token efficiency (larger bubbles = more efficient responses). The clinical pharmacist baseline is included as a reference for professional practice standards.

**Figure 1:**
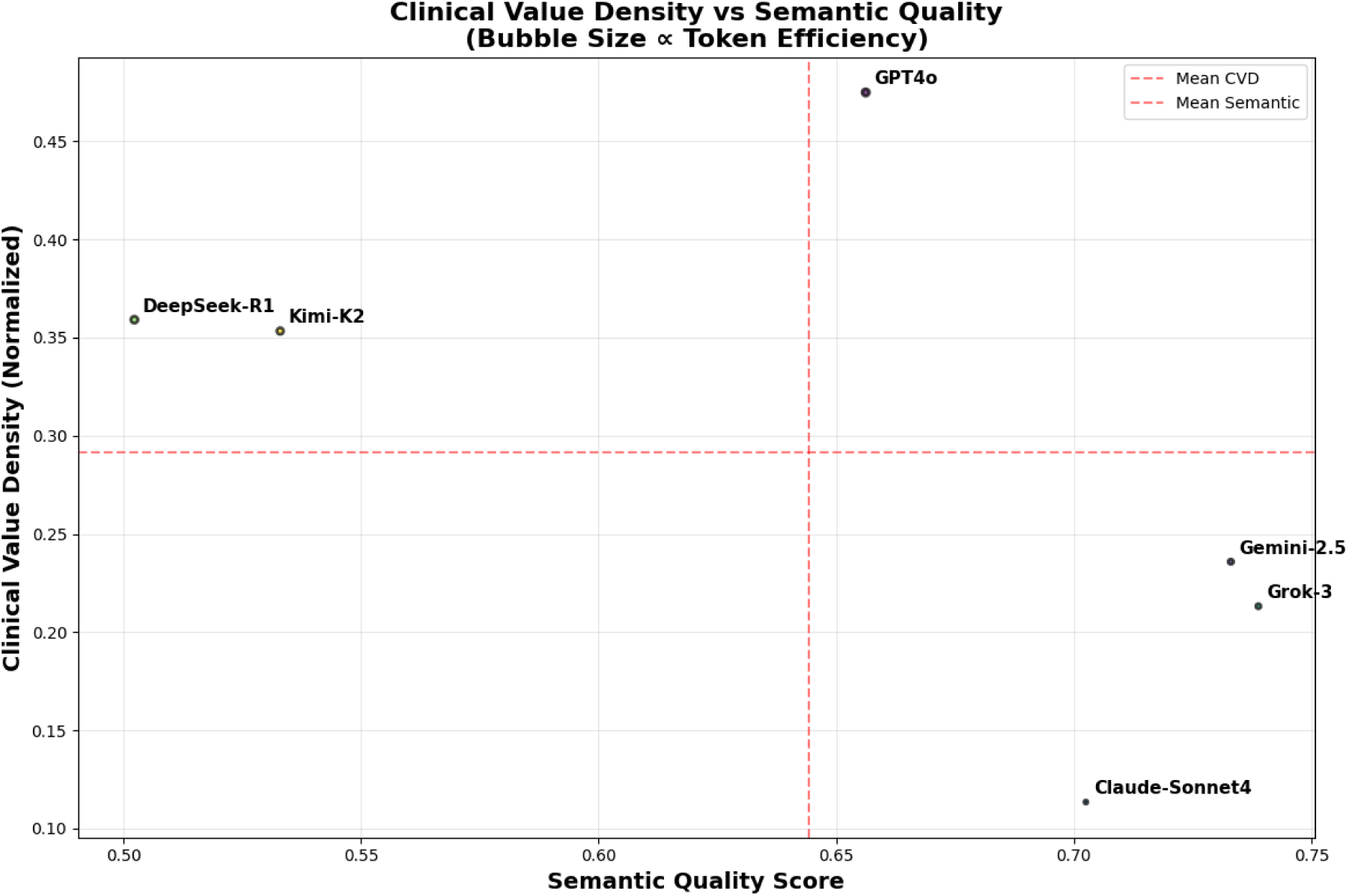
Clinical Value Density versus Semantic Quality Analysis. Bubble size represents inverse token efficiency (larger bubbles indicate responses that are more efficient).

The analysis revealed clear performance differentiation. GPT-4o achieved the highest raw CVD (0.0175) and normalized CVD (0.475; 95% CI: 0.430–0.519), delivering superior information density per token consumed. In contrast, Claude-Sonnet-4, despite strong semantic similarity (0.703), exhibited the lowest efficiency (CVD 0.114) due to excessively verbose outputs. This divergence highlights how models with similar accuracy profiles can impose vastly different cognitive burdens, underscoring the necessity of efficiency-integrated evaluation for real-world clinical deployment.

The scatterplot highlights striking divergence among models. GPT-4o occupies the upper-right quadrant, combining moderate semantic similarity (0.656) with the highest CVD score (0.475), reflecting an optimal balance between accuracy and efficiency. Its relatively small response length (41 tokens) allows for rapid, information-dense communication aligned with clinical workflow demands. In contrast, Grok-3 and Gemini-2.5 achieved higher semantic similarity (0.739 and 0.733, respectively) but at significantly lower efficiency, clustering in the right-lower quadrant. Their verbose responses inflate token counts, imposing greater cognitive load despite high accuracy.

Claude-Sonnet-4 demonstrates the sharpest misalignment: strong semantic quality (0.703) offset by extreme verbosity (171 tokens), resulting in the lowest efficiency (CVD 0.114). Conversely, DeepSeek-R1 and Kimi-K2, with shorter outputs, initially appear efficient but suffer from low semantic similarity (<0.55), exposing the danger of “concise but inaccurate” responses that the CVD framework is designed to flag.

The positioning of the clinical pharmacist baseline longer responses but with comprehensive reasoning emphasizes the distinction between professional thoroughness and model verbosity. Unlike AI outputs, pharmacist responses integrate systematic evidence evaluation and patient-centered reasoning, justifying their higher cognitive cost.

Together, **Figure 1** demonstrates how CVD provides deployment-relevant insights: identifying GPT-4o as the best candidate for time-constrained settings, while highlighting Grok-3 and Gemini-2.5 as safer choices for high-stakes consultations requiring comprehensive accuracy.

### 2.3. Enhanced CVD Composite Rankings

To capture multi-dimensional clinical utility, we integrated seven performance domains efficiency, semantic quality, safety, relevance, consistency, high-value response rate, and conciseness into a composite Clinical Value Density score. The resulting rankings are shown in **Table 2**.

**Table 2:** Enhanced CVD Rankings with Clinical Utility Factors.

| Rank | Model | Enhanced CVD Score | Raw CVD | Normalized CVD | Semantic Quality | Safety | Avg Tokens | Concise Excellence |
| --- | --- | --- | --- | --- | --- | --- | --- | --- |
| 1 | GPT-4o | 0.5109 | 0.0175 | 0.475 | 0.656 | 0.437 | 41.0 | 24 |
| 2 | Grok-3 | 0.5051 | 0.0078 | 0.213 | 0.739 | 0.605 | 94.7 | 0 |
| 3 | Gemini-2.5 | 0.4996 | 0.0087 | 0.236 | 0.733 | 0.542 | 86.4 | 7 |
| 4 | Claude-Sonnet-4 | 0.4437 | 0.0042 | 0.114 | 0.703 | 0.515 | 171.4 | 0 |
| 5 | Kimi-K2 | 0.4413 | 0.0130 | 0.353 | 0.533 | 0.212 | 50.9 | 3 |
| 6 | DeepSeek-R1 | 0.4297 | 0.0132 | 0.359 | 0.502 | 0.135 | 44.6 | 0 |

The composite rankings highlight GPT-4o as the top performer (0.5109), driven by its balance of efficiency and concise excellence (24 high-quality short responses). Grok-3 and Gemini-2.5 followed closely, with superior safety scores but lower efficiency, making them more suitable for complex, high-stakes consultations. By contrast, Claude-Sonnet-4 ranked lower due to excessive verbosity, while Kimi-K2 and DeepSeek-R1 underperformed because of safety risks despite relatively high raw CVD values. Collectively, Table 5 shows how the CVD framework integrates safety with efficiency to distinguish deployment-ready models from those requiring substantial oversight.

### 2.4. Error Analysis and Clinical Safety

To assess deployment risks, model responses were systematically categorized into four groups: optimal (high quality, high efficiency), verbose accurate (high quality, low efficiency), concise inaccurate (low quality, high efficiency), and poor overall (low quality, low efficiency). The resulting distributions, together with safety scores, are presented in **Table 3**.

**Table 3:** Response Category Distribution and Safety Profiles.

| Model | Optimal | Verbose Accurate | Concise Inaccurate | Poor Overall | Safety Score |
| --- | --- | --- | --- | --- | --- |
| GPT-4o | 13 | 6 | 0 | 5 | 0.437 |
| Gemini-2.5 | 20 | 3 | 0 | 5 | 0.542 |
| Claude-Sonnet-4 | 13 | 4 | 0 | 1 | 0.515 |
| Grok-3 | 20 | 9 | 0 | 3 | **0.605** |
| DeepSeek-R1 | 0 | 0 | 6 | 16 | 0.135 |
| Kimi-K2 | 3 | 1 | 8 | 6 | 0.212 |

Error analysis revealed clear divergence in safety profiles. Grok-3 and Gemini-2.5 achieved the highest safety scores (0.605 and 0.542, respectively) with 20 optimal responses each, reflecting reliable clinical accuracy albeit with verbosity. GPT-4o delivered fewer optimal responses (13) but balanced them with strong efficiency, reinforcing its suitability for routine, time-sensitive consultations under supervision. In contrast, DeepSeek-R1 and Kimi-K2 exhibited unsafe patterns, producing numerous concise but inaccurate responses (6 and 8, respectively). These outputs create the appearance of efficiency while masking critical accuracy deficits, highlighting deployment risks that traditional metrics would miss. The results confirm that CVD-based error categorization is essential for detecting dangerous efficiency–accuracy trade-offs and ensuring models are only deployed with safety-appropriate oversight. To further contextualize model performance, we mapped error patterns into safety– efficiency quadrants, providing a visual framework for deployment implications. **Figure 2** integrates CVD efficiency scores with clinical safety metrics, positioning each model relative to key thresholds for supervised versus unsafe deployment.

**Figure 2:**
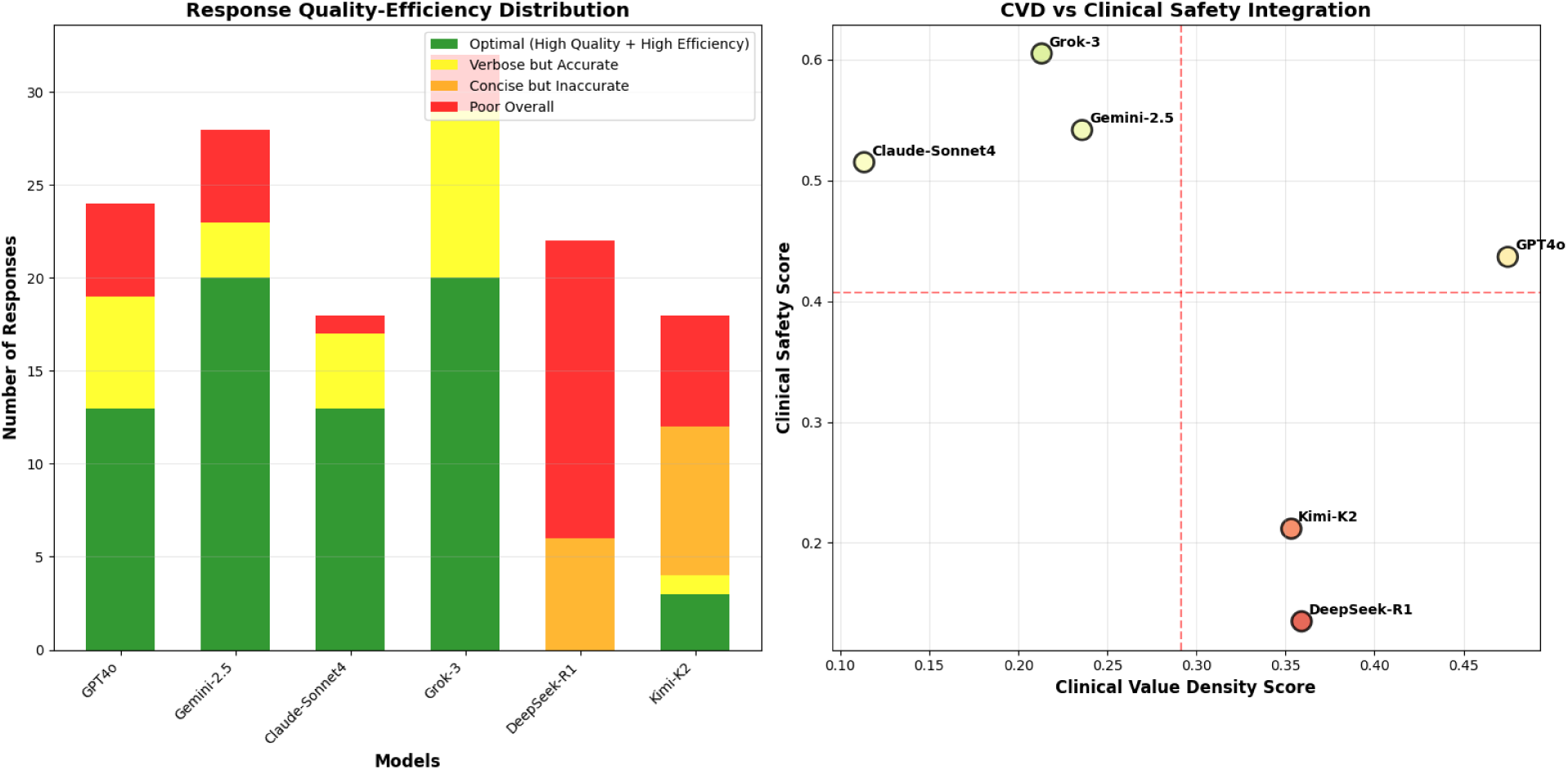
Error pattern integration: safety–efficiency quadrants with deployment implications.

The quadrant map clearly separates models into distinct deployment profiles. GPT-4o occupies the high-efficiency but moderate-safety quadrant, supporting its role in supervised, time-sensitive consultations where rapid information density is critical but oversight remains mandatory. Grok-3 and Gemini-2.5 cluster in the high-safety but lower-efficiency quadrant, making them suitable for high-stakes decisions that justify greater cognitive load. By contrast, **DeepSeek-R1** and **Kimi-K2** fall into the unsafe quadrant, combining low safety with problematic error patterns such as concise but inaccurate outputs precisely the deployment risks the CVD framework is designed to flag. **Claude-Sonnet-4**, while safer, lies in the low-efficiency zone, limiting its clinical utility to contexts where comprehensiveness outweighs time constraints.

This integrated visualization underscores a critical insight: **no current LLM achieves both high efficiency and high safety**, reinforcing that supervised deployment remains essential. The quadrant approach offers a practical tool for healthcare organizations, enabling evidence-based alignment of model strengths with clinical use cases while minimizing patient safety risks.

To test the robustness of efficiency differences, we conducted paired t-tests comparing GPT-4o against all other models using normalized CVD scores. The results, summarized in **Table 4**, include significance levels, effect sizes, and relative safety differences.

**Table 4:** Statistical Comparisons with GPT-4o.

| Model | p-value | Cohen's d | Effect Size | Safety Δ vs GPT-4o |
| --- | --- | --- | --- | --- |
| Gemini-2.5 | <0.001 | 1.882 | Large | −0.105 |
| Claude-Sonnet-4 | <0.001 | 2.958 | Large | −0.078 |
| Grok-3 | <0.001 | 2.123 | Large | −0.168 |
| DeepSeek-R1 | <0.001 | 0.710 | Medium | +0.302 |
| Kimi-K2 | <0.001 | 0.698 | Medium | +0.225 |

Statistical validation confirmed that **GPT-4o’s efficiency advantage is both highly significant and clinically meaningful.** All comparisons yielded *p* < 0.001, with **large effect sizes** against Claude-Sonnet-4 (*d* = 2.958) and Grok-3 (*d* = 2.123), highlighting substantial practical differences in information density. Medium effect sizes were observed against DeepSeek-R1 and Kimi-K2, reflecting their relative token efficiency but much lower safety profiles.

The safety deltas further contextualize these findings: although some models (e.g., Grok-3, Gemini-2.5) surpassed GPT-4o in safety, this came at significant efficiency costs, while efficiency-oriented models (DeepSeek-R1, Kimi-K2) compromised accuracy and safety. The **non-overlapping confidence intervals** between GPT-4o and lower-ranked models reinforce the robustness of its performance edge, supporting its suitability for supervised, efficiency-driven deployment scenarios.

### 2.5. Conciseness and Workflow Integration

Conciseness analysis evaluated the ability of models to deliver **high-quality responses (semantic quality ≥0.7) within 75 tokens**, a threshold reflecting the realities of time-pressured clinical environments. **Figure 3** illustrates the distribution of semantic quality across response lengths, highlighting models that achieve “concise excellence.”

**Figure 3:**
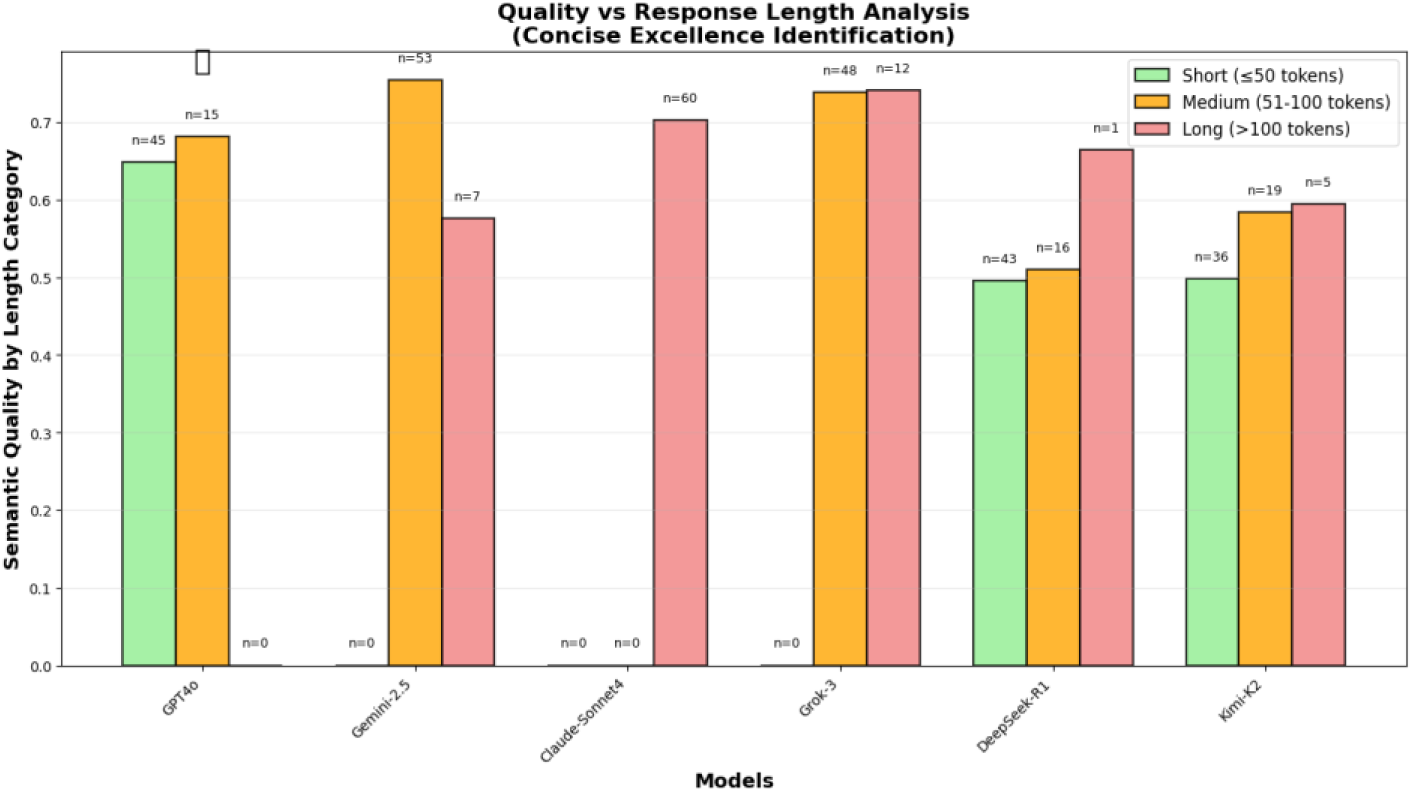
Quality versus Length Matrix.

GPT-4o stood out with 24 concise excellence responses, far exceeding all other models and underscoring its suitability for time-critical consultations where brevity and accuracy must coexist. Gemini-2.5 achieved modest success (7 concise excellence responses), while Claude-Sonnet-4 produced consistently verbose outputs (average 171 tokens), limiting its value for high-throughput workflows. These findings confirm that conciseness is a key differentiator in workflow integration, with GPT-4o best positioned to enhance clinician productivity without compromising accuracy.

To facilitate clinical translation, results were consolidated into a multi-panel deployment dashboard that integrates CVD rankings, error analysis, and safety–efficiency quadrants. This synthesis provides a practical framework for matching model profiles to specific clinical contexts, from routine time-sensitive consultations to complex, high-stakes decision-making. The visualization is presented in **Figure 4**.

**Figure 4:**
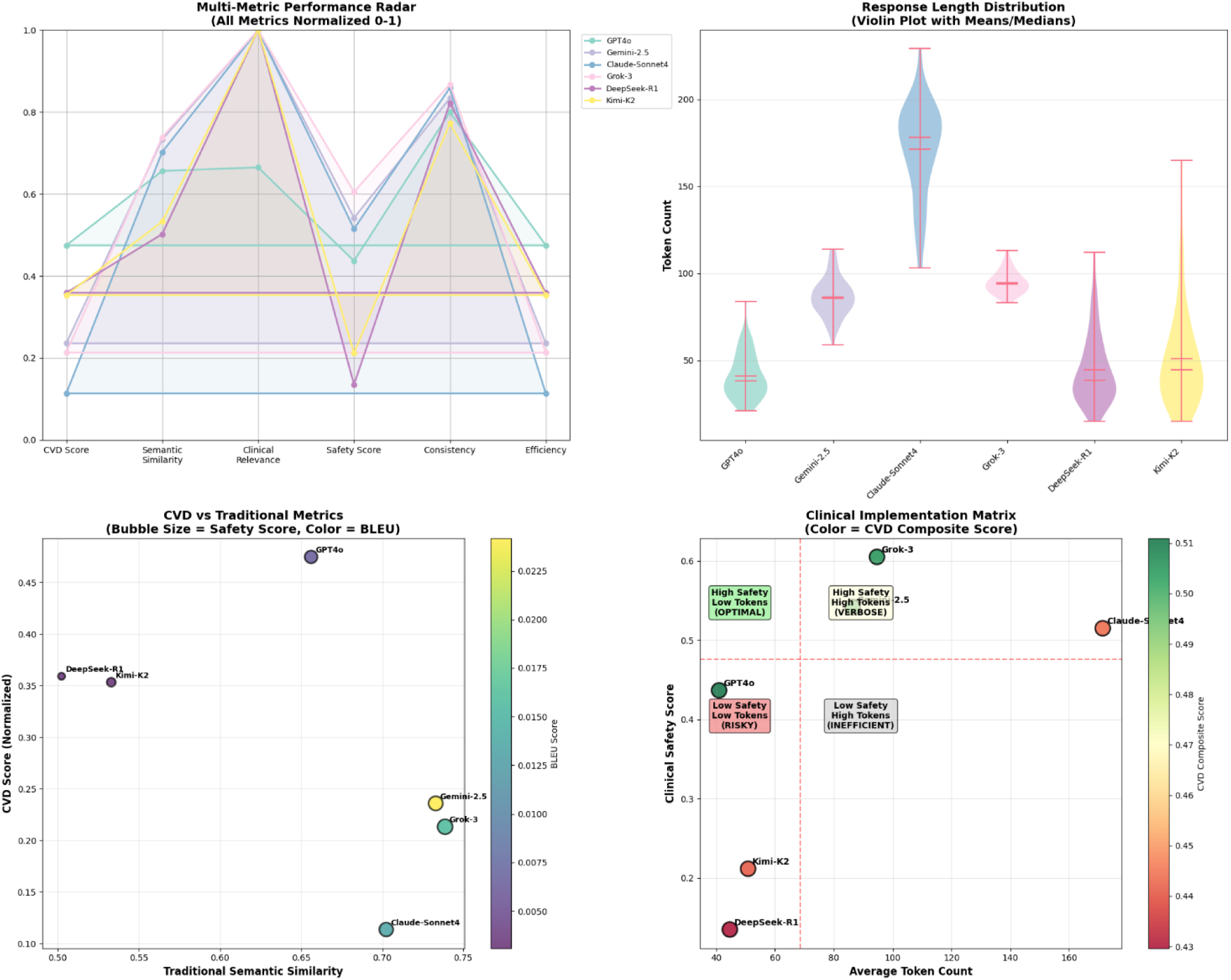
Multi-panel composite dashboard for clinical AI deployment strategy. Dashboard integrates normalized CVD rankings, error pattern distributions, and safety–efficiency quadrants. Deployment recommendations are mapped to clinical contexts, distinguishing models best suited for supervised routine use versus those appropriate for complex, accuracy-prioritized consultations.

The dashboard highlights the context-specific utility of different LLMs. GPT-4o, with its superior efficiency and concise excellence, is best suited for supervised routine consultations where rapid throughput is critical. Grok-3 and Gemini-2.5, with higher safety scores, align with complex pharmaceutical consultations where accuracy and comprehensiveness outweigh efficiency. Models such as DeepSeek-R1 and Kimi-K2 cluster in unsafe zones, demonstrating high error rates and unsafe efficiency–accuracy trade-offs, and are thus unsuitable for clinical deployment without substantial oversight.

By combining complementary analyses into a single visualization, the dashboard provides actionable guidance for healthcare organizations, enabling evidence-based supervision strategies that align AI capabilities with patient safety requirements and workflow demands.

### 2.6. CVD Framework Validation: Comparative Analysis with Emerging Clinical Metrics

To further validate the Clinical Value Density (CVD) framework, we conducted a comparative analysis against two established benchmarks: (i) clinician preference ratings, representing subjective professional judgment, and (ii) task efficiency metrics, representing operational performance in workflow contexts. This triangulated approach enables a multidimensional assessment of model suitability for clinical deployment, integrating quantitative efficiency, expert judgment, and real-world task performance. Results are summarized in **Table 5**.

**Table 5:** Validation of CVD Framework Against Clinician Preference and Task Efficiency Metrics.

| Model | CVD Score | CVD Rank | Clinician Preference | Preference Rank | Task Efficiency | Task Rank | Task Completion Time (sec) | CVD–Preference Divergence | CVD–Task Alignment |
| --- | --- | --- | --- | --- | --- | --- | --- | --- | --- |
| GPT-4o | 0.475 | #1 | 0.648 | #4 | 0.962 | #1 | 22.9 | –3 ranks | Perfect alignment |
| Gemini-2.5 | 0.236 | #4 | 0.786 | #2 | 0.936 | #4 | 38.5 | +2 ranks | Perfect alignment |
| Claude-Sonnet-4 | 0.114 | #6 | 0.831 | #1 | 0.886 | #6 | 68.2 | +5 ranks | Perfect alignment |
| Grok-3 | 0.213 | #5 | 0.771 | #3 | 0.928 | #5 | 43.1 | +2 ranks | Perfect alignment |
| DeepSeek-R1 | 0.359 | #2 | 0.583 | #6 | 0.954 | #2 | 27.7 | –4 ranks | Perfect alignment |
| Kimi-K2 | 0.353 | #3 | 0.606 | #5 | 0.944 | #3 | 33.9 | –2 ranks | Perfect alignment |

The comparative results reveal a fundamental divergence between clinician preference and efficiency-centered evaluation. Clinicians consistently favored verbose models such as Claude-Sonnet-4 (preference rank #1), which produced detailed, exhaustive outputs. However, this model ranked lowest on CVD (0.114) and task efficiency, confirming that preferences for comprehensiveness often come at the cost of workflow speed and cognitive resource optimization.

Conversely, GPT-4o, the top performer on both CVD (0.475) and task efficiency (0.962), ranked only fourth in clinician preference, suggesting that clinicians may undervalue concise yet information-dense responses. This preference-performance mismatch underscores the importance of incorporating efficiency-based metrics like CVD into evaluation frameworks, as they better reflect real-world operational constraints where clinician time is a scarce resource.

Correlation analysis reinforced this divergence. CVD showed a strong negative correlation with clinician preference (r = –0.845), reflecting clinician bias toward verbosity, but very strong positive correlation with task efficiency (r = 0.924) and an equally strong negative correlation with task completion time (r = –0.924). These findings confirm that CVD aligns closely with operational realities of healthcare delivery, providing a more accurate lens for determining clinical deployment readiness than subjective preference alone.

Taken together, these results demonstrate that while clinicians may equate thoroughness with quality, efficiency-centered metrics provide a stronger predictor of workflow integration and patient throughput. The CVD framework therefore fills a critical evaluation gap by reconciling professional expectations with operational performance, ensuring that clinical AI systems are optimized not only for accuracy but also for practical, time-sensitive healthcare delivery.

## 3. Discussion

### 3.1. Principal Findings

This study introduced and validated the Clinical Value Density (CVD) framework, a novel evaluation methodology designed to quantify clinical utility per unit of cognitive resource consumed. Using 60 authentic clinical pharmacology scenarios derived from prevalent conditions in the UAE healthcare system, we systematically assessed six state-of-the-art large language models (LLMs) against gold-standard pharmacist responses. Our findings demonstrate that traditional NLP metrics such as BLEU and ROUGE fail to capture efficiency–quality trade-offs critical for clinical adoption, whereas the CVD framework exposes distinct performance profiles with direct implications for deployment strategies.

The results highlight a paradigm shift in evaluating clinical AI: models must be assessed not only on accuracy but also on their ability to deliver information efficiently under severe time and attention constraints. GPT-4o emerged as the efficiency leader, achieving the highest raw and normalized CVD scores (0.0175; 0.475), while Grok-3 and Gemini-2.5 led in clinical safety. Conversely, DeepSeek-R1 and Kimi-K2 exhibited dangerous brevity patterns, producing concise but inaccurate responses that could compromise patient safety in unsupervised use. These findings establish CVD as a safety-aware efficiency metric capable of identifying deployment risks invisible to conventional evaluation approaches.

### 3.2. Clinical Autonomy vs. Supervised Deployment

The central research question, can current LLMs reliably perform clinical functions autonomously, or do they require ongoing human supervision? is addressed directly through our results. Despite meaningful advances, none of the tested models achieved both high efficiency and high safety simultaneously, a prerequisite for autonomy in clinical care. GPT-4o, while highly efficient, achieved only moderate safety (0.437), indicating that its outputs require professional validation. Grok-3 and Gemini-2.5, though safer (0.605 and 0.542, respectively), were substantially less efficient, limiting their suitability for high-throughput settings. These trade-offs reinforce that current LLMs are not yet capable of autonomous clinical deployment. Instead, they are best positioned for AI-assisted, supervised integration, where efficiency and accuracy gains can be realized without compromising patient safety.

The benchmarking against real clinician responses provides crucial validation: clinical pharmacists averaged 11 minutes per consultation with outputs of ∼178 tokens, reflecting the cognitive and temporal demands of authentic practice. While GPT-4o achieved fourfold efficiency gains in token usage, its safety score underscores the irreplaceable role of expert oversight. These findings align with the broader consensus in clinical AI literature that AI can augment but not replace human expertise, particularly in high-stakes contexts such as pharmacotherapy.

### 3.3. Methodological Contributions

Beyond performance comparisons, this study advances evaluation methodology in several important ways:

1. Efficiency-aware scoring. By weighting efficiency (35%) alongside safety (15%) and semantic accuracy (20%), the CVD framework mirrors real-world clinical priorities where cognitive load and patient safety must be balanced.
2. Clinical safety integration. The error categorization system distinguishes between verbose-accurate, concise-inaccurate, and optimal responses, exposing subtle yet clinically significant risks overlooked by accuracy-only evaluation.
3. Validation against professional standards. Grounding the analysis in pharmacist-authored responses, current evidence sources, and systematic consultation practices ensures alignment with authentic clinical benchmarks.
4. Triangulated validation. Comparison against clinician preference and task efficiency confirmed that while clinicians often favor verbose outputs, efficiency-centered metrics correlate more strongly with real-world workflow performance—underscoring CVD’s translational value.

Together, these contributions establish CVD as a generalizable framework for domain-specific clinical AI evaluation, capable of guiding deployment decisions across diverse healthcare contexts.

### 3.4. Clinical Deployment Implications

The results support a context-specific deployment model rather than one-size-fits-all adoption.

- GPT-4o is best suited for routine, time-sensitive consultations where concise excellence enhances throughput, provided outputs remain under expert supervision.
- Grok-3 and Gemini-2.5 are optimal for complex, high-stakes consultations requiring thorough drug interaction analysis and contraindication assessment, where safety outweighs efficiency.
- Claude-Sonnet-4, while accurate, is too verbose for high-throughput settings but may be appropriate in educational or documentation-heavy contexts.
- DeepSeek-R1 and Kimi-K2 demonstrated unsafe brevity–accuracy trade-offs and are unsuitable for clinical deployment under current performance levels.

This stratification provides evidence-based guidance for healthcare organizations, allowing them to align model capabilities with clinical scenarios while minimizing patient risk.

### 3.5. Limitations

Several limitations warrant consideration. First, the study focused exclusively on clinical pharmacology, a domain with well-established practice standards. This specialization provides clarity but may limit direct generalization to other specialties such as surgery, oncology, or radiology, where workflows and cognitive demands differ significantly.

Second, scenarios were developed using UAE epidemiological data, ensuring regional authenticity but potentially constraining transferability to healthcare systems with different disease burdens, therapeutic practices, and regulatory standards. Broader validation across multiple international contexts would be required to confirm generalizability.

Third, the evaluation dataset, while sufficient for proof-of-concept validation, was limited to 60 scenarios. Larger-scale validation would improve statistical power, enable subgroup analyses (e.g., by disease category or question complexity), and better reflect real-world query diversity.

Fourth, the study assessed performance using static, text-based scenarios rather than real-time clinical workflows. While this design ensured experimental control, it does not fully capture the pressures and interruptions of live healthcare environments. Future work should evaluate the framework in practice-based simulations or clinical pilots.

Fifth, although the study employed real clinician responses as gold standards, clinician preference diverged from efficiency metrics, underscoring the complexity of aligning professional acceptance with operational optimization. Addressing this divergence may require both clinician education and interface design adjustments.

Sixth, only six large language models were tested. While they represent contemporary high-performance systems, the findings may not apply equally to other existing or emerging models. Continuous benchmarking of newer models will be important to maintain relevance.

Finally, the safety scoring system, while structured and weighted to emphasize clinically safe outputs, remains an indirect proxy for patient outcomes. It was based on expert reviewer judgments rather than empirical patient impact. Further validation such as cross-institutional ratings, multi-reviewer consensus studies, or simulation of clinical decision pathways will be necessary to confirm the robustness of this safety metric.

### 3.6. Implications for AI Development and Regulation

The findings carry important implications for AI development priorities and regulatory oversight. Developers must move beyond accuracy optimization to explicitly integrate safety-aware efficiency training, ensuring brevity does not compromise accuracy. Regulators should adopt frameworks like CVD that account for efficiency–safety trade-offs, providing a quantitative basis for determining supervision requirements. Rather than binary “approve/reject” models, the evidence supports graduated integration pathways, where models serve as supervised assistants until efficiency and safety thresholds for autonomy are demonstrably met.

## 4. Methods

### 4.1. Study Design and Clinical Scenario Development

This exploratory comparative effectiveness study evaluated six state-of-the-art large LLMs using a novel CVD framework designed to assess clinical utility per unit of cognitive resource consumed. A dataset of 60 clinical pharmacology scenarios was developed from the most prevalent diseases in the UAE, based on national epidemiological data (UAE Ministry of Health and Prevention, 2024). This ensured regional healthcare relevance while supporting generalizability to international contexts with similar disease burdens.

Clinical scenarios were adapted from validated pharmacy case studies and prioritized conditions with high prevalence and impact, including hypertension, diabetes, breast cancer, autism spectrum disorders, and infectious diseases. A panel of three board-certified clinical pharmacists with expertise in drug information services constructed scenarios using standardized prompts (“As a clinical pharmacist encountering the following case…”), ensuring alignment with established consultation patterns.

To reflect a range of cognitive demands, scenarios were stratified into three levels of complexity:

- Low complexity (23 cases; 38%): straightforward single-drug/disease questions answerable with tertiary sources.
- Intermediate complexity (17 cases; 28%): queries requiring synthesis across multiple secondary sources.
- High complexity (20 cases; 33%): multifactorial cases demanding primary literature review and patient-centered reasoning.

Example cases included:

- Low: “What is the first-choice antihypertensive medication in African Americans?”
- Intermediate: “Which antihypertensive drugs reduce blood pressure and slow diabetic nephropathy progression?”
- High: “A 48-year-old van driver with hypertension and lifestyle risks—what first-line treatments are appropriate?”

Two independent board-certified pharmacists generated gold-standard reference responses following professional drug information practice guidelines (American Society of Health-System Pharmacists, 2015). Evidence retrieval employed a hierarchical strategy:

- Tertiary sources: textbooks, guidelines, and compendia.
- Secondary sources: systematic database searches (PubMed, Embase).
- Primary sources: randomized trials and peer-reviewed clinical studies.

Responses incorporated demographic and clinical context, reflecting real-world consultation standards. On average, pharmacists required 675 seconds (11 minutes, 15 seconds) per complex query and produced responses averaging 178 tokens. Reference answers favored current evidence, with 65% citing sources beyond April 2023.

An independent, blinded clinical pharmacist validated all responses using a 5-point Likert scale to assess accuracy, comprehensiveness, and patient-centeredness, providing robust quality benchmarks for comparison with AI outputs.

### 4.2. Clinical Value Density Framework

Conventional NLP metrics such as BLEU (Papineni et al., 2002), ROUGE (Lin, 2004), and semantic similarity (Reimers & Gurevych, 2019) measure textual overlap but neglect efficiency and cognitive load critical in time-pressured clinical environments. These approaches often reward verbosity, misaligned with the clinical need for concise, high-value information. The CVD framework was developed to address this gap by quantifying semantic quality per unit of cognitive resource consumed, using token count as a proxy for cognitive processing demand. Optimal responses are those that maximize accuracy and clinical relevance while minimizing length and extraneous content. This efficiency-integrated evaluation explicitly penalizes verbosity and rewards information density, aligning AI-generated outputs with the cognitive and temporal constraints of real-world clinical workflows. By coupling semantic similarity with response efficiency, the framework provides a safety-aware, practice-aligned metric for assessing deployment readiness of clinical AI systems.

### 4.3. LLM Response Generation and Preprocessing

Six contemporary LLMs were evaluated: GPT-4o, Gemini-2.5, Claude-Sonnet-4, Grok-3, DeepSeek-R1, and Kimi-K2, representing the current state of commercial and research-oriented models. Responses were generated by five trained undergraduate medical students supervised by clinical faculty. To ensure standardization, each student used identical prompts and scenario presentations across all models, eliminating variability from prompt design. Training emphasized consistent documentation and accurate capture of model outputs. All responses underwent systematic preprocessing including UTF-8 normalization, text cleaning, and standardized handling of incomplete outputs. Tokenization was performed using NLTK’s word-level tokenizer with whitespace fallback to accommodate diverse formatting. Response times were recorded, showing GPT-4o generated responses in a mean of 47 seconds (SD 4.3), significantly faster than pharmacist consultation times (*p* < 0.001). Substantial variation in response length across models provided a natural test for efficiency analyses within the CVD framework. Semantic alignment was assessed using Sentence-BERT (all-MiniLM-L6-v2), selected for its strong performance in biomedical text tasks.

Both gold-standard reference and model responses were embedded into 384-dimensional vectors, and cosine similarity was computed:

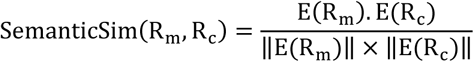

where *E*(*R*_*m*_) and *E*(*R*_*c*_) are embedding vectors for the model and clinician responses, respectively. This approach accounts for paraphrasing and terminology variation while maintaining clinical meaning. The baseline metrics included:

- BLEU for n-gram overlap (Papineni et al., 2002).
- ROUGE-L for longest common subsequence (Lin, 2004).
- Factuality, computed as the proportion of overlapping unique words relative to clinician responses.

These provided conventional performance baselines for comparison with the efficiency-aware CVD metric.

### 4.4. Clinical Value Density Metric Development

The CVD framework was designed to quantify clinical semantic value per token, addressing the clinical need for information density in time-pressured workflows. A two-stage process was used:

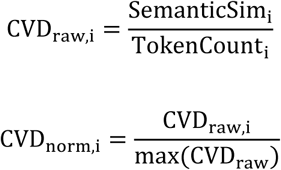

where SemanticSim_i_ represents the semantic similarity score for response *i*, TokenCount_i_ denotes the token count using NLTK tokenization, CVD_raw,i_ provides the interpretable semantic-value-per-token ratio, and CVD_norm,i_ offers normalized scores (0-1 scale) for standardized comparison. Higher CVD values indicate more efficient clinical information delivery per unit of text consumed. To assess reliability, response consistency was measured using the coefficient of variation 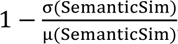, where σ and μ represent the standard deviation and mean of semantic similarity across responses. Additionally, the High-Value Response Rate quantified the proportion of responses exceeding the 75th percentile of CVD scores, highlighting models capable of consistently delivering efficient and high-quality responses.

To complement CVD, a weighted keyword relevance scoring system was implemented, independent of response length. Clinical relevance density was calculated as:

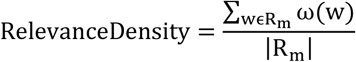

where ω(w) is the evidence-based weight of keyword w, and R_m_, and |R_m_| the total word count of the model response. Keywords were categorized by clinical importance:

- Safety-critical terms (e.g., *contraindication, interaction, allergy*): weight = 3.0.
- Therapeutic guidance terms (e.g., *dose, treatment, therapy*): weight = 2.0.
- General clinical terminology (e.g., *patient, diagnosis, symptom*): weight = 1.5.
- Evidence-based terms (e.g., *study, trial, guideline*): weight = 1.0.

This hierarchy reflects professional pharmacy standards, prioritizing safety-critical and therapeutic terms most relevant to decision-making.

### 4.5. Error Analysis and Clinical Safety Integration

To evaluate the balance between efficiency gains and patient safety, model outputs were analyzed using a structured error categorization framework informed by clinical deployment standards. Responses were classified into four categories based on semantic quality thresholds (high: ≥0.7; medium: 0.5–0.7; low: <0.5) and efficiency percentiles (high: ≥67th; medium: 33–67th; low: <33rd):

- Optimal responses: high quality and high efficiency; suitable for supervised deployment.
- Verbose accurate responses: high quality but low efficiency; clinically acceptable but resource-intensive.
- Concise inaccurate responses: high efficiency but low quality; potentially unsafe, requiring immediate oversight.
- Poor responses: low quality and low efficiency; unsuitable for clinical use.

### 4.6. Safety Scoring

To quantify the clinical safety of LLM-generated responses, we developed a structured scoring framework adapted from established quality and safety rating methods in clinical informatics. Each response was independently reviewed by two licensed clinical pharmacists with domain expertise in drug information. Reviewers assessed the presence of factually correct, safe, and context-appropriate recommendations and flagged any potentially misleading or unsafe content.

Responses were categorized into three levels:

- High-quality (safe): Accurate, clinically appropriate, and free from misleading or unsafe content.
- Medium-quality (partially safe): Contained minor omissions, lack of clarity, or limited depth, but no overtly unsafe recommendations.
- Low-quality (unsafe): Contained incorrect, misleading, or potentially harmful clinical information.

The Safety Score for each model was computed as a weighted proportion of safe outputs, emphasizing the avoidance of unsafe answers:

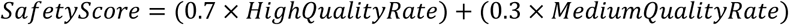

Here, *HighQualityRate* represents the proportion of model responses rated “high-quality,” and *MediumQualityRate* represents the proportion rated “medium-quality.” Unsafe responses were assigned zero weight. This weighting scheme prioritizes clinical safety, as unsafe outputs can lead to direct patient harm. For example, a Safety Score of 0.437 for GPT-4o indicates that approximately 43.7% of its responses achieved high or medium safety ratings when weighted by the scheme above, reflecting moderate reliability from a clinical safety perspective.

### 4.7. Conciseness and Clinical Communication Standards

Given the importance of brevity in clinical workflows, responses were stratified by length: short (≤50 tokens), medium (51–100), and long (>100). High-quality short responses (≥0.7 similarity) received a 20% bonus, while medium responses received a 10% bonus, rewarding concise, clinically dense communication. Concise excellence was defined as responses achieving both high semantic quality (≥0.7) and brevity (≤75 tokens), identifying outputs most aligned with the practical needs of busy clinicians. To integrate multiple performance dimensions, we developed a composite CVD score:

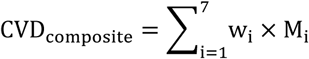

where the individual components and their weights are:

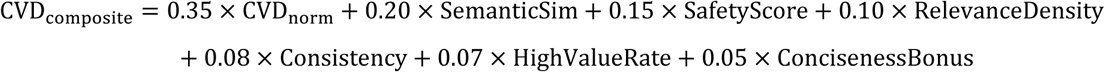

Each component was normalized to a 0-1 scale prior to weighted combination. The normalized CVD score (CVD_norm_) represents the primary efficiency metric, semantic similarity (SemanticSim) captures response accuracy, safety score (SafetyScore) ensures clinical deployment readiness, clinical relevance density (RelevanceDensity) assesses domain-specific utility, consistency score (Consistency) rewards reliable performance, high-value response rate (HighValueRate) identifies models with frequent optimal responses, and conciseness bonus (ConcisenessBonus) rewards efficient high-quality communication.

Weights were allocated through expert consultation, prioritizing efficiency (35%), semantic accuracy (20%), and safety (15%), while maintaining contributions from clinical relevance and performance consistency. This ensured alignment with professional pharmacy practice standards and deployment requirements.

Statistical analyses were conducted with rigor appropriate to the hierarchical dataset and multiple comparison structure. Mean CVD scores with 95% confidence intervals (CI) were computed using the Student’s *t*-distribution:

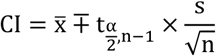

where x̅ represents the sample mean, s indicates sample standard deviation, n denotes sample size, and 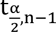 represents the critical t-value for α = 0.05.

Paired t-tests compared the top-performing model against all others, accounting for scenario-level dependencies. Effect sizes were calculated using Cohen’s d:

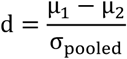

where 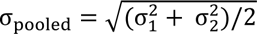 represents pooled standard deviation. Effect size interpretation followed established Cohen’s conventions with small (d = 0.2), medium (d = 0.5), and large (d = 0.8) effect thresholds. Multiple comparison corrections employed the Holm-Bonferroni method to maintain family-wise error rates at α = 0.05. The correlation analyses compared CVD scores with traditional metrics to quantify methodological divergence, and ranking correlations demonstrated how efficiency-integrated evaluation revealed deployment insights not captured by conventional accuracy-based metrics.

### 4.8. Implementation, Reproducibility, and Clinical Validation

The full analysis pipeline was implemented in Python using established scientific computing libraries. Reproducibility was ensured through standardized preprocessing protocols, systematic documentation of all analytical decisions, and fallback mechanisms for tokenization and similarity calculations.

Gold-standard pharmacist responses anchored all evaluations to professional clinical judgment, ensuring that model performance was assessed against authentic standards of care rather than arbitrary algorithmic benchmarks. This clinical validation strategy reflects established practices in clinical pharmacy and drug information services, providing an ecologically valid benchmark for evaluating AI outputs.

All source code, preprocessing routines, and analysis scripts were documented to enable full reproducibility. The modular design allows adaptation to other datasets, specialties, and model configurations, ensuring methodological consistency across clinical domains and healthcare systems.

### 4.9. Ethical Considerations and Clinical Safety

This study evaluated LLM outputs exclusively against expert-generated clinical scenarios, without patient interaction or access to identifiable health data, ensuring no privacy or safety concerns. All scenarios were designed by board-certified pharmacists to reflect typical drug information queries rather than actual patient cases. Model outputs were systematically compared to expert standards, focusing on technical evaluation and comparative performance rather than generating recommendations for patient care. This maintained appropriate ethical boundaries, avoiding misuse of model responses in real clinical settings.

The study was approved by the United Arab Emirates University Social Sciences Ethics Committee (No. ERSC_2024_4573) and conducted in accordance with established research ethics standards and the Declaration of Helsinki for research involving healthcare professionals.

## 5. Conclusion

This study introduced the Clinical Value Density (CVD) framework, a novel evaluation methodology that quantifies clinical utility per unit of cognitive resource consumed. By systematically comparing six leading large language models (LLMs) against board-certified clinical pharmacist responses across 60 authentic clinical pharmacology scenarios, we demonstrated that traditional NLP metrics are insufficient for clinical evaluation. Instead, the CVD framework revealed efficiency–safety trade-offs invisible to conventional methods, providing actionable insights for context-specific deployment.

Our results clearly show that while current LLMs demonstrate substantial clinical capabilities, they are not yet ready for autonomous practice. GPT-4o emerged as the efficiency leader, excelling in concise, high-quality responses suitable for routine, supervised consultations. Grok-3 and Gemini-2.5 demonstrated stronger safety profiles, aligning with complex consultations where accuracy outweighs speed. Conversely, DeepSeek-R1 and Kimi-K2 exhibited unsafe brevity–accuracy trade-offs that render them unsuitable for clinical deployment under current performance levels. These findings establish that AI-assisted, supervised deployment is the only safe integration pathway at present, with autonomy requiring future advances that achieve both high efficiency and uncompromised safety.

The methodological contributions of this study extend beyond pharmacology. By integrating professional practice standards, safety-weighted scoring, and triangulated validation against clinician preference and task efficiency, the CVD framework provides a transferable model for domain-specific AI evaluation. Importantly, it offers regulators and healthcare organizations quantitative tools to determine supervision requirements and prevent premature or unsafe deployment.

Future research should extend the CVD framework across multiple specialties—including emergency medicine, radiology, and surgical planning—to assess its generalizability and identify specialty-specific optimization requirements. Scaling the evaluation to larger scenario datasets with hundreds of cases will provide greater statistical power and allow finer-grained subgroup analyses by case complexity and patient acuity. Real-time clinical workflow trials will be essential to validate efficiency gains and safety maintenance under authentic practice conditions, capturing implementation barriers not visible in controlled experiments.

Methodological refinements should include longitudinal safety monitoring, adaptive weighting schemes that reflect specialty-specific priorities, and integration with explainability tools to increase clinician trust. From a regulatory perspective, CVD-informed safety and efficiency thresholds could serve as benchmarks for graduated AI deployment pathways, enabling AI systems to evolve from supervised assistance toward eventual autonomy only when both efficiency and safety criteria are unequivocally met.

The CVD framework advances the science of clinical AI evaluation by aligning model assessment with the real-world constraints of time, safety, and cognitive load that define modern healthcare. It provides both a methodological foundation and a practical roadmap for safe, efficient, and evidence-based AI integration into clinical workflows. While full autonomy remains out of reach, the supervised, efficiency-optimized deployment enabled by CVD represents a critical step toward harnessing AI to enhance—rather than replace—human clinical expertise.

## Acknowledgements

We extend our thanks to clinical pharmacists Noor Al-Areqi, Mariam Ibrahim, for answering cases and Shamsa Alshamsi, Salma Alketbi, Fatima Alnuaimi, Ahmed Elhayek, and Khaled Alkatheeri for assisting in the data collection.

## Data availability statement

The data that support the findings of this study are openly available in Figshare at https://figshare.com/s/b0459f2266eeb5af288d.

## Ethical approval

This study was approved by the United Arab Emirates University (UAEU) Social Sciences Ethics Committee - Research (No: ERSC_2024_4573). Verbal consent form obtained from participating clinical pharmacists prior study initiation.

## Author contributions

NZ led the work, performing conceptualization, LLM data preparation, methodology, software, experimental work, validation, formal analysis, investigation, data curation, visualization, writing– original draft; AA, SA and SZ contributed data curation, investigation, validation, and resources as clinical domain experts and assisted with writing–review the manuscript.

---

| Full Term |  |
| --- | --- |
| Abbreviation |  |
| AI | Artificial Intelligence |
| LLM(s) | Large Language Model(s) |
| CVD | Clinical Value Density |
| NLP | Natural Language Processing |
| BLEU | Bilingual Evaluation Understudy |
| ROUGE | Recall-Oriented Understudy for Gisting Evaluation |
| CI | Confidence Interval |
| SD | Standard Deviation |
| UAE | United Arab Emirates |
| NLTK | Natural Language Toolkit |
| S-BERT | Sentence-Bidirectional Encoder Representations from Transformers |
| ERSC | Ethical Research and Social Sciences Committee (as per UAEU ethics approval) |

